# Statistical analysis plan for a stepped-wedge cluster randomized clinical trial for the evaluation of the clinical impact of different telemedicine practices in intensive care units – TELESCOPE II trial

**DOI:** 10.64898/2026.01.13.26344052

**Authors:** Tiago Mendonça Dos Santos, Renato Carneiro de Freitas Chaves, Bruna Gomes Barbeiro, Maura Cristina Dos Santos, Thiago Domingos Corrêa, Alexandre Biasi Cavalcanti, Ary Serpa Neto, Carlos Henrique Sartorato Pedrotti, Fernando Zampieri, Guilherme de Paula Pinto Schettino, Jorge Ibrain Figueira Salluh, Leandro Utino Taniguchi, Leonardo José Rolim Ferraz, Luciano Cesar Azevedo, Otávio Berwanger, Regis Goulart Rosa, Renata Albaladejo Morbeck, Rodrigo Biondi, Suzana Margareth Lobo, Jessica Kasza, Adriano José Pereira, Otavio Ranzani

**Author notes:** **Corresponding author**: Tiago Mendonça dos Santos Hospital Israelita Albert Einstein Department of Critical Care Medicine Avenida Albert Einstein, 627/701, 5° floor Zip code: 05651-901 - São Paulo (SP), Brazil. TMS and RCFC have contributed equally to this study.

## Abstract

**Background:** The optimal model for delivering Tele-ICU is still to be determined. Previous clinical trials focused on medical-led daily multidisciplinary rounds (DMRs), showing certain improvements in processes of care but without direct evidence for improvement in clinical outcomes.

**Objective:** To evaluate whether a structured Tele-ICU intervention, comprising DMRs led by an intensivist plus a multidisciplinary care bundle (nursing, physical therapy, pharmacy) and a quality and safety management package, can reduce ICU length of stay in Brazilian public ICUs.

**Methods:** The TELESCOPE 2 is a multicenter, open-label, stepped-wedge cluster randomized controlled trial including 25 ICUs in Brazil from January 2024 to January 2026. In a stepped-wedge assignment, ICUs are randomized and allocated into one of five sequences. All adult patients admitted in participant ICUs will be eligible for inclusion in the study. Admissions for non-medical reasons, and patients previously included in the TELESCOPE 2 study will be excluded. All ICUs will receive the interventions staggered at different times. The trial intervention is multifaceted, comprising three components delivered in combination via telemedicine: i) daily multidisciplinary rounds led by board-certified intensive care physician; ii) coordinated care by a multidisciplinary team, including nurses, physiotherapists, and clinical pharmacists; iii) a management strategy focused on quality improvement and patient safety. The primary outcome is ICU length of stay. Secondary outcomes include clinical endpoints (ICU mortality, ventilator-free days at 28 days, and ventilator-associated events), process-of-care indicators (e.g., mobilization density, head-of-bed elevation, and sedation management), and unit-level organizational outcomes based on standard resource use and standardized mortality rates.

**Conclusion:** We describe the statistical analysis plan for the TELESCOPE II trial, finalized prior to database lock. This pre-specified approach mitigates analysis bias and ensures the transparency and robustness of the reported results.

**TRIAL REGISTRATION:** Clinicaltrials NCT05960994, Brazilian Registry of Clinical Trials RBR-342wxn9, and Universal Trial Number U1111-1298-9799.

## INTRODUCTION

Tele-ICU is a technology-enabled model of care that allows intensivists to remotely support bedside teams. This approach has gained traction as a response to the uneven distribution of intensivists and limited access to critical care in underserved regions of Brazil, as in several other countries.(1)

Tele-ICU interventions, such as remote daily multidisciplinary rounds (DMRs), real-time clinical support, and performance monitoring, have been associated with improvements in patient care, including decrease in ICU mortality, shorter hospital length of stay, and faster responses to alarms. Nevertheless, the best delivery model of Tele-ICU is uncertain.(1) The two randomized clinical trials (ERIC and TELESCOPE-I trials)(2,3) focused on ICU settings could not show direct improvements in clinical outcomes, although improvement in certain processes of care was observed.(2) One of the potential reasons for the lack of success on showing benefit in clinical outcomes was the model of delivery, i.e., intensivist-centered, without specifically accounting for the clinical multidisciplinary team and organizational factors. While the feasibility and benefits of tele-ICU are well documented, there is limited evidence on integrating telemedicine with multidisciplinary care and quality management strategies.

The TELESCOPE-II study aims to evaluate whether a structured telemedicine intervention, comprising DMRs led by an intensivist, a multidisciplinary care bundle (nursing, physical therapy, pharmacy), and a quality and safety management package, can reduce ICU length of stay in Brazilian public ICUs.

## Methods

### Study design

The TELESCOPE-II study is an open-label, national, multicenter, stepped-wedge cluster randomized controlled trial. The unit of randomization is the intensive care unit (ICU), as the intervention targets the entire multidisciplinary team. A total of 25 ICUs will be randomized into five sequences (**Figure 1**). A complete list of participating centers and trial committee members is provided in the Supplementary Appendix.

**Figure 1:**
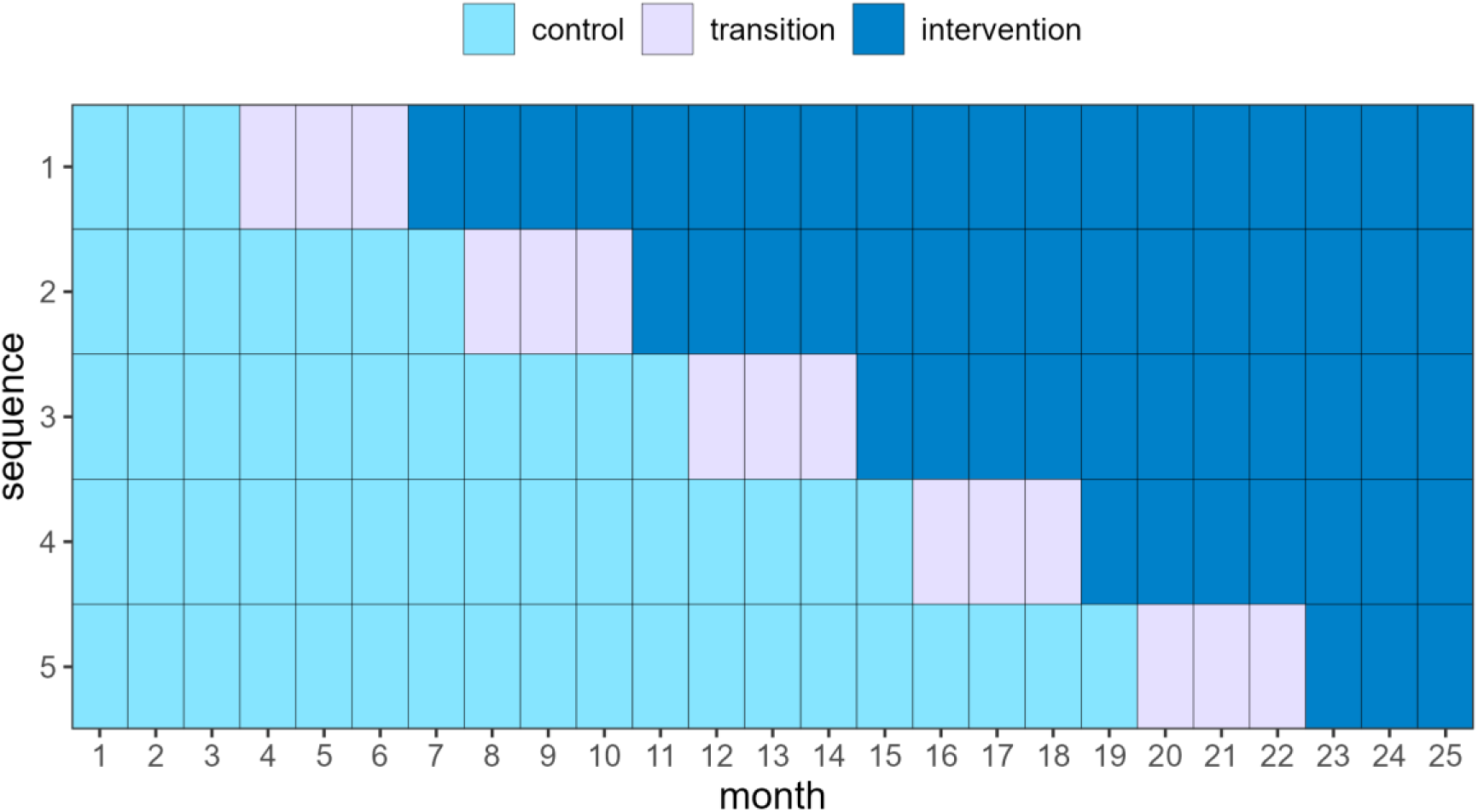
Schematic diagram illustrating the trial timeline. Five ICUs are randomized to each sequence.

Each sequence consists of a control period without any intervention, followed by a staggered implementation of the intervention according to the assigned sequence. Given the complexity of the intervention package, a delay in the full effect of the intervention package being attained is anticipated. Therefore, all sequences include a transition period during which the intervention implementation will commence, but data collected during the transition period will be excluded from the primary analysis. All ICUs will eventually receive the full intervention package at different time points, enabling both within- and between-sequence comparisons.

### Randomization and Allocation

Centers will be randomized into five sequences using a stratified approach. Randomization will be performed using preliminary data from the first two months of the baseline period. The first baseline period will consist of 3 months; however, the third month will be designated for organizational preparation and disclosure of allocation for the first sequence.

The stratification will be based on two variables: geographic region and ICU length of stay (LOS).

- Region (**Figure 2**)

`o` South: centers located in the South and Southeast regions
`o` North: centers located in the Central-West, North, and Northeast regions
- ICU Length of Stay (LOS):

`o` Below: centers whose median ICU length of stay is lower than the median for their respective region
`o` Above: centers whose median ICU length of stay is higher than the median for their respective region

**Figure 2:**
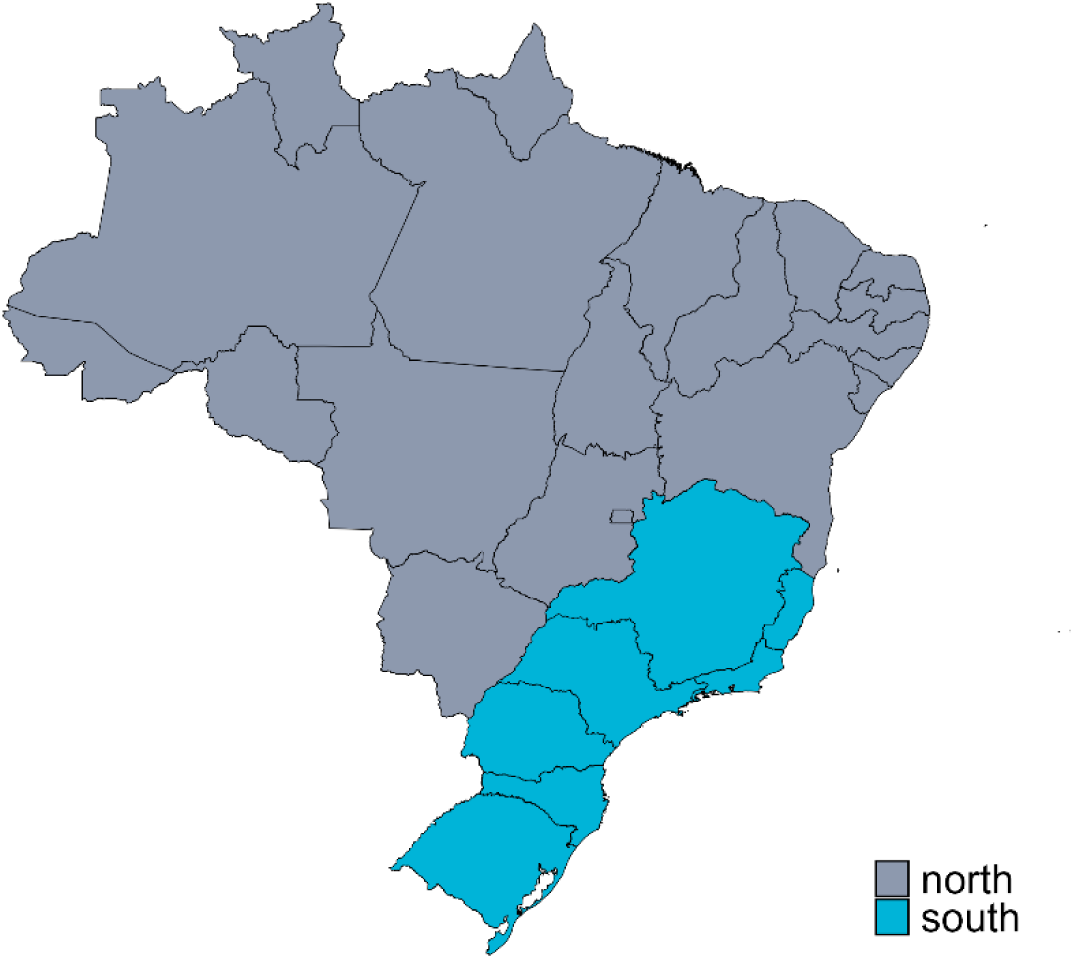
Regional Grouping of Brazilian States for Randomization Purposes.

This stratification, generating four strata, was adopted to enhance balance across sequences, as geographic region in Brazil is associated with several structural and healthcare-related characteristics.(3,4) The stratum combining the South region and below-median LOS included 7 centers, while the other three strata included 6 centers each.

All 25 ICUs will be randomized simultaneously using a computer-generated allocation list prepared by an independent statistician. The randomization will be constrained to a subset of sequences to ensure regional balance and approximate balance in baseline ICU LOS across sequences (and within strata) to deal with the uneven number of centers by stratum. Specifically, sequences 1, 3, and 5 will be constructed to include one additional center from the South region, while sequences 2 and 4 will included one additional center from the North region. The exact distribution of centers across the four strata (Region and ICU LOS) for each sequence is detailed in Table 1. Candidate allocations not meeting the region and baseline LOS constraints will be excluded, and one allocation will be selected uniformly at random among the eligible allocations. This constrained randomization will preserve overall balance while maintaining the probabilistic nature of the allocation.(5)

**Table 1:** Distribution of centers by sequence according to stratification variables (Region and ICU LOS).

| Sequence | North / Below median LOS | North / Above median LOS | South / Below median LOS | South / Above median LOS | Total centers |
| --- | --- | --- | --- | --- | --- |
| 1 | 1 | 1 | 2 | 1 | 5 |
| 2 | 1 | 2 | 1 | 1 | 5 |
| 3 | 1 | 1 | 1 | 2 | 5 |
| 4 | 2 | 1 | 1 | 1 | 5 |
| 5 | 1 | 1 | 2 | 1 | 5 |
| **Total** | 6 | 6 | 7 | 6 | 25 |

Allocation concealment will be ensured by restricting access to the randomization sequence to the statistician, who will only have access to ICU identification codes and will be blinded to unit identities. The allocation list will be subsequently provided to the study coordinator, who will disclose each center’s assignment 30 days prior to the transition phase to allow for organizational preparation.

The randomization will be performed using R (version 4.2.2).(6) Primary analyses will adjust for region and baseline LOS category, thereby acknowledging the constrained randomization.

### Sample size calculation

The required sample size was determined through simulation of a mixed-effects model for ICU length of stay based on 1,000 replications for each scenario and using Kenward-Roger correction. (7) The model included fixed effects for the intercept (2.079), a time trend (-0.012 i.e. a decrease in log-length of stay of 0.012 units per month), and the intervention effect (-0.148), corresponding to a 13.75% reduction from a baseline mean of 8 days (i.e., a reduction of 1.1 days). The random effects were characterized by: cluster-level variability following a normal distribution with a variance of 0.102; a period-within-cluster effect following a normal distribution with a variance of 0.019; and an individual-level residual error following a normal distribution with a variance of 1.090. These values yield an estimated intracluster correlation coefficient (ICC) of 0.10 and a cluster autocorrelation coefficient (CAC) of 0.84. These empirical estimates were derived from the TELESCOPE-I study.(3) Based on these simulations, a total sample size between 18,750 and 25,000 patients is required to achieve at least 95% power at a two-sided significance level of 5%. This range reflects an expected monthly recruitment rate of 30 to 40 patients per ICU.

Because center withdrawal is a potential contingency in multicenter studies, we assessed the robustness of the design under two scenarios: removal of three centers from the first sequence and removal of three centers from the last sequence. Under a conservative scenario with 30 patients per ICU per period and removal of three centers from any sequence, power remains at least 93%.

### Interim analysis

Since our intervention incorporates the best available evidence for the care of critically ill patients in ICUs, and no inherent risks are anticipated in the trial’s execution, interim analyses are not planned. As a result, the formation of a formal data monitoring committee was deemed unnecessary.

#### Eligibility criteria

The selected public and philanthropic ICUs were invited via electronic communication to participate in an interview to assess the inclusion and exclusion criteria, using an electronic feasibility assessment questionnaire. All patients admitted to the included ICUs will be eligible for inclusion in the study. The inclusion and exclusion criteria for both ICUs and eligible patients are detailed below.

#### Inclusion criteria for ICUs

Public or philanthropic hospitals with 7 to 20 beds; availability of physicians and nurses 24 hours a day and a physiotherapist available for at least 18 hours a day.

#### Exclusion criteria for ICUs

(i) ICUs with structured multidisciplinary round, defined as meetings (DMRs) ≥ 3 times per week, during weekdays, conducted by a certified intensivist and documented in medical records with fixed visit length (>5 min / patient), using some supporting tool (checklist or standard form), goal-oriented, based on established protocols, including all the patients admitted to the ICU; or (ii) ICUs with implemented monthly management of indicators (audit and feedback) with specific planning; or (iii) coronary or cardiac intensive care units, or other specialized units (e.g., neurological, or burn units); or (iv) step-down units or intermediate care units; or (v) ICUs without renal replacement therapy availability; or (vi) ICUs in which coordinators are board-certified intensivists and qualified as Masters of Business Administration (or an equivalent).

#### Inclusion criteria for patients

Adults (≥ 18 years old).

#### Exclusion criteria for patients

Admissions for non-medical reasons (e.g., judicial) and previously included in the Telescope 2 study.

## Outcomes

The follow-up period to define all outcomes will be truncated at 90 days while in the hospital from ICU admission.

### Primary outcome

The primary outcome of this trial at the patient level is ICU length of stay (LOS) defined as the time interval in hours between patients’ ICU admission and the moment of ICU physical discharge (i.e., transfer to another care facility or another hospital) or ICU death, as defined by the hospital’s system date and time. Date and time will be entered by the health care worker responsible for data collection. ICU LOS will be derived in 24 hours periods with decimal places, as recommended.(8)

### Secondary outcomes

- ICU mortality, defined as death by any cause during the index ICU admission.
- In-hospital mortality, defined as death by any cause during the index hospital admission.
- Ventilator-free days during the first 28 days, defined as the number of days alive and free from mechanical ventilation for at least 24 consecutive hours from successfully weaning to day 28. Patients who die before weaning will be deemed to have zero ventilator-free days, and patients discharged from the hospital before 28 days will be considered alive and free from mechanical ventilation at 28 days.
- ICU readmission within 48 hours.(9)
- Early reintubation (<48h after elective extubation).
- Ventilator-associated events: defined as either an increase in the daily minimum positive end-expiratory pressure (PEEP) of ≥3 cmH2O sustained for at least two consecutive calendar days following a period of at least two days of stable or decreasing PEEP, or an increase in the fraction of inspired oxygen (FiO2) by ≥20 percentage points sustained for at least two consecutive days after a minimum of two days of stable or decreasing FiO2 levels.(10)
- Accidental extubation rate.

#### Process-of-care and quality indicators

- Patient mobilization density in the ICU: defined as the number of days the patient undergoes any form of mobilization, including passive movement of upper or lower limbs in bed, sitting on the edge of the bed or in a chair, or walking.
- Adherence to maintaining the head-of-bed elevation (30°-45°).
- Adequate prevention of venous thromboembolism. (11)
- Rate of patient-days under adequate sedation [defined by Richmond agitation and sedation scale (RASS) = -3 to +1]. (12)
- Rate of patients-days with oral or enteral nutrition.
- Rate of patients with adequate glycemic control (defined as blood glucose between 70 to 180mg/dL). (3, 11)
- Rate of patients-days within normoxemia (defined as peripheral oxygen saturation = 92 to 96%). (3)
- Rate of central venous catheter use.
- Central venous catheter dwell time.
- Rate of indwelling urinary catheter use.
- Indwelling urinary catheter dwell time.

#### Unit-level organizational outcomes

- ICU classification by standard resource use (SRU) and standardized mortality rate (SMR): defined by standard resource use (SRU) and standardized mortality rate (SMR).(13) SRU reflects the observed-to-expected rate of resource utilization, estimated as ICU length of stay (LOS) for surviving patients and adjusted for the patient’s severity of illness. SMR reflects the observed / expected rate, according to acute physiology score (SAPS3) of hospital deaths. The profiles are a combination of SMR (above or below median) and SRU (above or below median). Each unit will be assigned to one of four groups: “most efficient” (SMR and SRU below median); “least efficient” (SMR, SRU above median); “overachieving” (low SMR, high SRU), “underachieving” (high SMR, low SRU).

### Statistical Principles

All statistical tests will be two-sided, and a significance level of 5% will be used for all analyses. Primary analyses will follow the intention-to-treat principle, with patients analyzed according to the randomized assignment of their ICU, regardless of intervention exposure (i.e. ICUs will be considered to have switched to the intervention at their scheduled time). The reporting of findings will follow the standards established by the CONSORT extension for stepped-wedge trials.(14)

Analyses will be conducted using R software (version 4.3.3 or later, Vienna, Austria). Mixed-effects models (described below) will primarily be fitted with the lme4 package.(15) For continuous outcomes, due to the reduced number of clusters, the Kenward-Roger small-sample correction will be applied. Prespecified secondary outcomes and subgroup analyses will not be adjusted for multiple comparisons and will be considered exploratory.

Multiple imputation will be performed if missing data on core variables exceed 10%, following standard procedures for multiple imputation using chained equations with mice package. (16)

### Primary outcome analysis

The primary outcome, ICU LOS, will be analyzed using a linear mixed-effects model applied to the natural logarithm of ICU LOS (in days). This transformation addresses the typically right-skewed distribution of LOS. The model will include both fixed and random effects to account for the hierarchical structure of the stepped-wedge design, the stratified randomization variables and patient level SAPS 3 score.

Formally, the model is defined as:

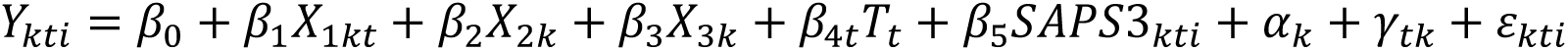

Where:

- 𝑌_𝑘𝑡𝑖_: log- transformed ICU LOS for patient *i* at time period *t* in cluster *k*;
- 𝛽_0_: fixed intercept (expected log-LOS at baseline);
- 𝛽_1_: fixed effect for the intervention;
- 𝑋_1𝑘𝑡_: intervention indicator for cluster *k* at time *t*, defined as 1 for intervention and 0 for control;
- 𝛽_2_: fixed effect for the stratification variable of Brazilian region;
- 𝑋_2𝑘_: regional location for cluster k, defined as 0 for North region centers and 1 for South region centers;
- 𝛽_3_: fixed effect for the stratification variable of ICU LOS category;
- 𝑋_3𝑘_: baseline ICU LOS category for cluster *k,* defined as 0 for below-median and 1 for above-median within region;
- 𝛽_4𝑡_: fixed effect for calendar time;
- 𝑇_𝑡_: factor for month;
- 𝛽_5_: fixed effect for SAPS3;
- 𝑆𝐴𝑃𝑆3_𝑘𝑡𝑖_: SAPS3 for patient *i* in period t and cluster k;
- 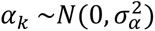: random intercept for cluster *k* (ICU), capturing between-cluster variability;
- 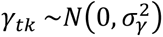: random effect for time within cluster, accounting for within-cluster temporal correlation;
- 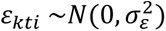: individual-level residual error.

This model accounts for both clustering and repeated measures over time within each ICU and allows estimation of the intervention effect while adjusting for temporal trends and between-cluster variability.

### Secondary, process-of-care, and unit-level organizational outcomes analysis

Continuous secondary and exploratory outcomes will be analyzed using the same mixed-effects modeling framework adopted for the primary outcome, including the same fixed and random effects. Count and rate outcomes will be analyzed using generalized linear mixed-effects models with Poisson or negative binomial distributions, as appropriate, incorporating the same fixed and random effects structure used in the primary model.

Binary outcomes will be analyzed using generalized linear mixed-effects models with a logit link function, incorporating the same set of independent variables used in the primary model. These models will account for clustering at the ICU level and repeated measures over time.

For categorical outcomes with more than two categories, multinomial logistic mixed-effects models will be fitted with ICU random intercepts specified at the cluster (ICU) level. The same covariate structure applied to the primary outcome model analysis will be used. These models will allow for estimation of category-specific odds ratios while accounting for hierarchical data structure.

### Sensitivity analysis for the primary outcome

We will assess the robustness of the primary analysis to alternative modeling choices and design assumptions as follows.

I. Covariate adjustments. We will repeat the primary analysis additionally adjusting for the following covariates: type of ICU admission (three categories: medical, elective surgical, unplanned surgical), invasive mechanical ventilation at ICU admission (two categories: yes, no), number of ICU beds at baseline (continuous term), and ICU performance at baseline (four categories: most efficient, least efficient, overachieving, underachieving). For baseline covariates, baseline will be defined using data from the first 2 months of the study.
II. Alternative correlation structures. We will assess the robustness of our results to different assumptions regarding the within-cluster correlation. This will include fitting a model considering the cluster as the only random effect and allowing for a discrete-time decay model to account for the potential weakening of correlation over time
III. Readiness for discharge. To minimize bias from non-clinical delays (e.g., weekends or ward-bed availability), we will redefine ICU LOS as time from ICU admission to the calendar day on which the patient first met readiness-for-discharge criteria. (3)
IV. Transition window. We will also perform an analysis treating the transition period itself as part of the intervention to account for potential effects occurring during this phase.
V. Competing risk. Because the primary endpoint, ICU length of stay, combines two processes (discharge alive and ICU death), death functions as a competing event for discharge. To assess robustness, we will reanalyze time to ICU discharge alive within a competing-risk framework that treats death as a competing event. We will present cause-specific hazard ratio estimates and cumulative incidence for discharge alive. (17)
VI. Early mortality exclusion. We will repeat the primary analysis after excluding patients who died within the first 24 hours of ICU admission. Because the intervention has little to no opportunity to affect outcomes among patients who are critically ill on arrival and die very early, this analysis assesses whether such early deaths dilute or distort the estimated intervention effect.

### Time-dynamic effects

We will include an interaction between intervention and time since implementation within ICU to assess potential ramp-up or waning of effect over time. For this exploratory analysis we will also include the transition period for each sequence. Following the exposure time indicator framework, the treatment effect will be reported as separate point treatment effects for each month since implementation. (18)

### Subgroup analysis

Prespecified subgroup analyses will be conducted by introducing interaction terms between the intervention and each subgroup variable in the primary outcome model. The following subgroups will be explored:

- Type of admission (medical vs surgical)
- Telemedicine experience (centers with vs. without prior telemedicine use)
- SAPS3 score (categorized by tertiles)
- Mechanical ventilation status at admission (invasive vs. non-invasive vs. none)
- ICU performance at baseline (four categories: most efficient, least efficient, overachieving, underachieving). Each classification will be made based on the first two months of the initial baseline period.

These analyses will be exploratory in nature. Estimates of interaction effects will be interpreted with caution, without adjustment for multiple comparisons.

### Handling of Timing in Stepped-Wedge Design

In this stepped-wedge cluster randomized trial, the timing of intervention rollout across ICUs is a key component of the analytical structure. Each ICU contributes data during control and intervention phases, separated by a transition period during which the intervention is being implemented but is not yet assumed to have reached full effect.

For the primary analysis, only patients admitted during the defined control and intervention periods will be included. Patients admitted during the transition period will be excluded, as this phase is expected to involve partial uptake and variability in adherence to the intervention protocols.

Calendar time will be modeled as a fixed effect in all primary and secondary analyses to adjust for potential secular trends and other time-related confounders. This approach helps isolate the intervention effect from temporal changes unrelated to the study.

By excluding transition periods from the main analysis and explicitly accounting for time in the statistical model, we aim to minimize bias due to gradual implementation and better estimate the true effect of the intervention.

### Funding

The Brazilian Ministry of Health (Institutional Development Program of the Unified Health System-PROADI SUS) was the primary source of funding, including costs of physician services, purchase of equipment (hardware) for Telemedicine sessions, hiring of local professionals for data collection and travel expenses for training and monitoring. The same funding also covered costs related to the regulatory part of the study-data collection, monitoring, data curation and statistical support. The Einstein Hospital Israelita allocated time of professionals and specialists who sat on the executive and steering committee of the study, as well as assigned its Telemedicine service system.

Jessica Kasza is supported by a National Health and Medical Research Council Investigator Grant (ID: 2033380).

## Data Availability

All data produced in the present study are available upon reasonable request to the authors.

## Acknowledgements

The authors would like to thank the central TELESCOPE 2 team, clinical and data collection teams of each participating ICU, as well as the Einstein Hospital Israelita, and the Brazilian Ministry of Health.

## Notes

### Competing Interest Statement

The authors have declared no competing interest.

### Clinical Trial

Clinicaltrials NCT05960994, Brazilian Registry of Clinical Trials RBR-342wxn9, and Universal Trial Number U1111-1298-9799.

### Author Declarations

The study was approved by local Research Ethics Committee of the coordinating study center (Einstein Hospital Israelita, CAAE: 69575123.0.1001.0071), as well as by local IRBs from each one of the 25 participant centers, in compliance with Brazilian legislation.

