## Additional File for "Statistical analysis plan for a stepped-wedge cluster randomized clinical trial for the evaluation of the clinical impact of different telemedicine practices in intensive care units – TELESCOPE II trial"

**LIST OF HOSPITALS PARTICIPATING IN THE STUDY.**

- Hospital Ana Nery, Salvador, BA.

- Hospital das Clínicas Luzia de Pinho Melo, Mogi das Cruzes, SP.

- Hospital de Emergência Dr. Daniel Houly, Arapiraca, AL.

- Hospital de Emergência e Trauma Dom Luiz Gonzaga Fernandes, Campina Grande, PB.

- Hospital Dr. Genesio Rego, São Luís, MA.

- Hospital Dr. Leo Orsi Bernardes, Itapetininga, SP.

- Hospital Mangabeira Governador Tarcísio Burity, João Pessoa, PB.

- Hospital Estadual de Doenças Tropicais Dr. Anuar Auad, Goiânia, GO.

- Hospital Estadual de Luziânia, Luziânia, GO.

- Hospital Geral de Roraima, Boa Vista, RR.

- Hospital Maternidade e Pronto Socorro Santa Lúcia, Poços de Caldas, MG.

- Hospital Municipal de Santarém, Santarém, PA.

- Hospital Municipal Padre Germano Lauck, Foz do Iguaçu, PR.

- Hospital Municipal Salgado Filho, Rio de Janeiro, RJ.

- Hospital Municipal Senhora Santana, Brasília de Minas, MG.

- Hospital Nossa Senhora das Dores, Itabira, MG.

- Hospital Padre Máximo, Venda Nova do Imigrante, ES

- Hospital Regional de Barbacena, Barbacena, MG.

- Hospital Regional De Cáceres Dr. Antônio Fontes, Cáceres, MT.

- Hospital Regional Justino Luz, Picos, PI.

- Hospital Região Leste (Paranoá), Brasília, DF

- Hospital São Lucas, Garça, SP.

- Hospital Tramandaí, Tramandaí, RS.

- Irmandade da Santa Casa de Misericórdia de Mococa, Mococa, SP.

- Santa Casa de Paranavaí, Paranavaí, PR.

**COLLABORATORS:**

***Coordinating Center*:** Hospital Israelita Albert Einstein (HIAE).

**Executive and steering committee**: Renato Carneiro de Freitas Chaves, Bruna Gomes Barbeiro, Maura Cristina dos Santos, Tiago Mendonça dos Santos, Thiago Domingos Corrêa, Otavio T. Ranzani, Adriano José Pereira.

**Advisory and scientific committee:** Alexandre Biasi Cavalcanti, Ary Serpa Neto, Carlos Henrique Sartorato Pedrotti, Fernando Zampieri, Guilherme de Paula Pinto Schettino, Jessica Kasza, Jorge Ibrain Figueira Salluh, Leandro Utino Taniguchi, Leonardo José Rolim Ferraz, Luciano Cesar Azevedo, Otávio Berwanger, Regis Goulart Rosa, Renata Albaladejo Morbeck, Rodrigo Biondi, Suzana Margareth Lobo.

***Experts Committee (multidisciplinar team):*** Alessandra Gomes Chauvin, Aline Cristina Pedroso, Barbara Barduchi, Beatriz Rocha Monteiro, Camila de Carvalho Gambin, Cilene Saghabi, Eliton Paulo Leite Lourenço, Erika Yumiko Kumoto, Fabiana Rossi Varallo, Fernanda Paulino Fernandes, Flavia Oliveira Rodrigues, Flavia Veronezi Stankevicius, Gabrielli Pare Guglielmi, Gean Carlos Alves Moraes, Giovana Roberta Zelezoglo, Lidiane Soares Sodre da Costa, Luciana Laversveiler Moraes da Costa, Jessica Tamiris Romano, Joao Paulo Victorino, Marcele Pessavento, Raquel Afonso Caserta Eid, Renata de Souza Cyrino, Roberta Gonsalez dos Santos, Silvana Maria de Almeida, Tatiana Aporta Marins.

***Experts Committee (management team):*** Amanda Valle, Ana Cláudia Ferraz, Ana Lúcia Martins da Silva, Bruno de Arruda Bravim, Bruno Mazza, Daiane Seger, Felipe Maia de Toledo Piza, Gilberto Friedman, Glauco Whestphal, Guilherme de Paula Pinto Schettino, Gustavo Faissol Janot de Matos, Haggéas da Silveira Fernandes, Hipólito Carraro Jr., Joan Castro, Juliana Anacleto, Marcele Pesavento, Murillo Santucci Cesar de Assunção, Nelson Akamine, Niklas Soderberg Campos, Walace de Souza Pimentel.

**Research back office:** Ana Cristina Lagoeiro Patrocinio da Cruz, Andrea de Carvalho, Lucelio De Sousa Rocha, Lenine Melo Lino, Rodrigo Flor de Moura.

**Funding:** The Brazilian Ministry of Health (Institutional Development Program of the Unified Health System-PROADI SUS) was the primary source of funding, including costs of physician services, purchase of equipment (hardware) for Telemedicine sessions, hiring of local professionals for data collection and travel expenses for training and monitoring. The same funding also covered costs related to the regulatory part of the study-data collection, monitoring, data curation and statistical support. The Einstein Hospital Israelita allocated time of professionals and specialists who sat on the executive and steering committee of the study, as well as assigned its Telemedicine service system.

Jessica Kasza is supported by a National Health and Medical Research Council Investigator Grant (ID: 2033380).
